# Evaluation of WHO listed COVID-19 qPCR primers and probe in silico with 375 SERS-CoV-2 full genome sequences

**DOI:** 10.1101/2020.04.22.20075697

**Authors:** Derek Toms, Julang Li, Hugh Y. Cai

## Abstract

Quantitative reverse-transcription PCR (qRT-PCR) assays remains the gold standard for detection of the SARS-CoV-2 virus because of its sensitivity and specificity. However, successful design of qRT-PCR assays requires accurate viral genome sequences. With mutations accumulating as the virus is transmitted globally, we sought to compare current assays recommended by the World Health Organization with available SARS-CoV-2 genomic sequences *in silico*. While most sequences were conserved, there were notable mismatches, particularly in assays developed using early sequences when compared to more recent isolates. We recommend that any assay being evaluated for diagnostic tests be compared with prevalent sequence data from the region of proposed testing and that continued publicly accessible sequence information continue to be provided by the research community.

## Introduction

During infection outbreak in crisis like the COVID-19 pandemic, diagnostics are a crucial step to manage the rate of infection, especially when clinical symptoms are difficult to distinguish from other respiratory infections such as influenza. Public health measures decisions, such as a patient and contact tracing requiring further quarantine and surveillance are intimately related to whether a suspected case has been confirmed. Therefore, speed and accuracy of such tests are paramount, thus the development and application of sensitive and reliable diagnostic tests are critical.

Among several platforms available, quantitative (real-time) reverse-transcription polymerase chain reaction (qRT-PCR) remains the primary means for diagnosing the novel coronavirus SARS-CoV-2, the pathogen responsible for COVID-19 (reviewed in 1). Using short DNA oligonucleotides (primers and probes) that are complementary to specific sequences of viral genetic material, qRT-PCR diagnostic tests thus are based on detection of the genetic material of the virus and require accurate design to ensure detection sensitivity and specificity^2,3^. Primers, one on each strand of DNA serve as starting points for the DNA polymerase enzyme that carries out the RT-PCR reaction, while probes bind between the primer sites and confer specificity. Design of primers and probes is based on sequenced viral genomes that have been publicly available since late December 2019 and typically target regions of the open reading frame (ORF) 1ab, envelope (E) and nucleocapsid (N) coding regions^4^.

One of our major challenges in the diagnosis of COVID-19 is that detection sensitivity and specificity of SARS-CoV-2 genetic material using qRT-PCR are variable and sometimes low^5^. Multiple factors may have contributed to the low sensitivity of SARS-CoV-2 detection: location of clinical sampling; low patient viral load; sporadic shedding; and variation in detection kits from different manufacturers. One of the key factors determining kit detection sensitivity is how efficiently primers and probes bind target genetic material. This in turn is dependent on kit manufacturers using the most appropriate viral genome sequence data. We hypothesized that mutations between SARS-CoV-2 isolates may cause imperfect binding of the primers and probes and may contribute to ongoing issues with qRT-PCR detection and sensitivity. To test our hypothesis, we performed a sequence alignment between 375 SARS-CoV-2 genomes available from the GenBank and probe-primer testing sets recommended by the World Health Organization (WHO).

## Results

We performed nine in silico evaluations of SARS-CoV-2 qRT-PCR primers and probes listed in the protocols published on the WHO website^6^. Summarized results are shown in Table 1 with detailed explanations below:

**Table 1.** Alignment between primer and probe sets to SARS-CoV-2 genomic sequences

| Assay source | Target coding region | Oligo | Mismatch (bp) | Isolate country | Accession |
| --- | --- | --- | --- | --- | --- |
| China CDC | ORF1ab | REV primer | 1 | China | LR757997 |
| China CDC | N | FOR primer | 3 | Spain | MT233522 |
| China CDC | N | FOR primer | 3 | Israel | MT276598 |
| China CDC | N | FOR primer | 3 | India | MT163714 |
| China CDC | N | FOR primer | 3 | Peru | MT263074 |
| China CDC | N | FOR primer | 3 | USA | MT246470 |
| China CDC | N | FOR primer | 3 | USA | MT276327 |
| China CDC | N | FOR primer | 3 | USA | MT276329 |
| China CDC | N | FOR primer | 3 | USA | MT276330 |
| China CDC | N | FOR primer | 3 | USA | MT263402 |
| China CDC | N | FOR primer | 3 | USA | MT258379 |
| China CDC | N | FOR primer | 3 | USA | MT259250 |
| China CDC | N | FOR primer | 3 | USA | MT259263 |
| China CDC | N | FOR primer | 3 | USA | MT246451 |
| China CDC | N | FOR primer | 1 | USA | MT263410 |
| China CDC | N | FOR primer | 1 | USA | MT246456 |
| China CDC | N | REV primer | 2 | USA | MT263411 |
| Institute Pasteur | RdRp | REV primer 2 | 1 | China | MT226610 |
| Institute Pasteur | RdRp | REV primer 4 | 1 | USA | MT259238 |
| Institute Pasteur | E | Probe | 1 | Korea | MT039890 |
| USA CDC | N | FOR primer 2 | 1 | USA | MT258382 |
| USA CDC | N | Probe 2 | 1 | USA | MT263435 |
| USA CDC | N | Probe 2 | 1 | USA | MT263458 |
| Japan | N | Probe | 1 | USA | MT159720 |
| Japan | N | FOR primer | 1 | Japan | LC534419 |
| Berlin | E | Probe | 1 | Korea | MT039890 |
| Hong Kong | ORF1ab | REV primer | 1 | Iran | MT163712 |
| Hong Kong | ORF1ab | REV primer | 1 | USA | MT276327 |
| Hong Kong | N | Probe | 1 | USA | MT263458 |
| Hong Kong | N | Probe | 1 | USA | MT263435 |
| Thailand | N | REV primer | 1<sup>a</sup> | USA | MT184913 |

<sup>a</sup>Sequence MT184913 had an ambiguous “Y” denoting C or T, which therefore has a mismatch likelihood of 50% with the primer.

China CDC protocols are designed to amplify SARS-CoV-2 ORF1ab and N regions. For the ORF1ab region, the forward primers and probe had 100% identity to the binding sites of 374/375 full genome sequences. The reverse primer had 1 bp mismatching with one full genome sequence. However, for N gene amplification, a three base pair mismatch was found in the first 3 bp at 5’ end of the forward primers with the target site in sequences of 13 reported SARS-CoV-2 across various countries outside China. They are one each from Spain, Israel, India and Peru, and 9 genome sequences from the USA (Fig. 1). This is significant, as a PCR reaction with a primer with 3 bp mismatch with the primer biding site may not be functional, i.e. the PCR may generate false negative results when applied to above mentioned SARS-CoV-2 infected samples. In addition, the forward primer has 1 mismatched with sequence MT263410 (SARS-CoV-2/human/USA/WA-UW330/2020) and MT246456 (SARS-CoV-2/human/USA/WA-UW199/2020), and the reverse primer 2 bp mismatch with sequence MT263411 (SARS-CoV-2/human/USA/WA-UW331/2020).

**Figure 1.**
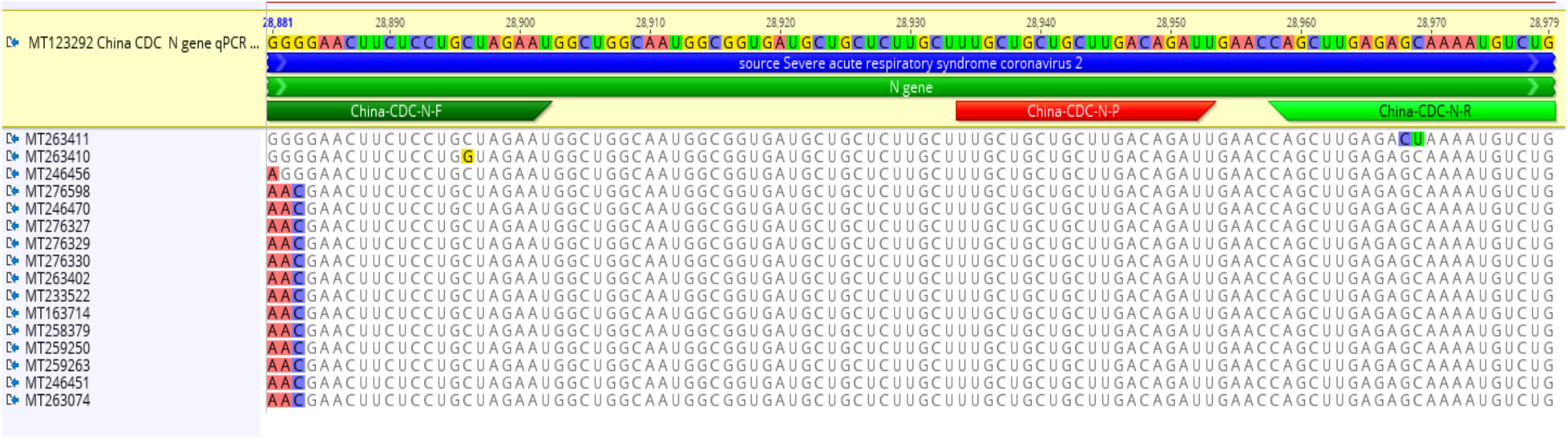
N gene primers of China CDC COVID-19 qRT-PCR had mismatches with multiple SARS-CoV-2 sequences. Note that primer and probe sequences are annotated on the top line (“MT123292 China CDC N gene qPCR”) while sequence mismatches with other isolates are highlighted below.

Next, we looked at qRT-PCR assays for the detection of SARS-CoV-2 from the Institut Pasteur, Paris. The primers and probe sequences were based on the first sequences of SARS-CoV-2 made available on the GISAID database on January 11, 2020. The primers and probes (nCoV_IP2 and nCoV_IP4) were designed to target the RNA-dependent RNA polymerase (RdRp) gene within the ORF1ab region spanning nt 12621-12727 and 14010-14116 (positions according SARS-CoV, NC_004718). The E gene assay from the Charité protocol was used as a confirmatory assay.

Institut Pasteur RdRp gene qRT-PCR primers and probe of both target sites (RdRp gene/nCoV_IP2 and RdRp gene/nCoV_IP4) have 100% identity to the binding sites of 374/375 full genome sequences. The reverse primer of the two PCR has 1 bp mismatching with sequence MT226610 (SARS-CoV-2/human/CHN/KMS1/2020) and MT259238 (SARS-CoV-2/human/USA/WA-UW246/2020), respectively. The primers and probe of the confirmation E gene qPCR had 100% identity with 376/377 SARS-CoV-2 E gene sequences available in GenBank. The probe had 1 bp mismatch with sequence MT039890 (isolate SNU01 from a Korean patient imported from Wuhan).

The current SARS-CoV-2 detection protocols provided by the US CDC includes two targets within the N gene. US CDC N gene 1 target primer and probe had 100% homology with 100% (387/387) available N gene (target 1) sequences of SARS-CoV-2 in GenBank. The N gene 2 target primers and probe are 100% homologous with 98.4% (379/385) N gene (target 2) of SARS-CoV-2 sequences. The probe had 1 bp mismatch with two sequences, i.e.MT263435 (SARS-CoV-2/human/USA/WA-UW355/2020) and MT263458 (SARS-CoV-2/human/USA/WA-UW379/2020). The forward primer had 1 bp mismatch with 1 sequence (MT258382, SARS-CoV-2/human/USA/CZB-RR057-014/2020).

Japan’s protocols included those for both gel PCR and qRT-PCR protocol. Here only the qRT-PCR primers and probe are evaluated in silico. The primers and probe had 100% identity with the binding site for 99.5% (384/386) of the available SARS-CoV-2 sequences. The probe had 1 bp mismatch with sequence MT159720 (2019-nCoV/USA-CruiseA-4/2020) and the forward primer had 1 bp mismatch with sequence LC534419 (SARS-CoV-2/Hu/Kng/19-437 RNA from Japan).

The protocol for diagnostic detection laid out in Berlin on January 17, 2020 consists of RdRp and E gene qRT-PCR using one probe (P2) specific for SARS-CoV-2 RdRp and a generic probe (P1) that cross-reacts with SARS-CoV and bat SARS-related CoVs, in addition to SARS-CoV-2. The other set of primers and probe is specific for the E gene of SARS-CoV-2. In Canada, Public Health Ontario uses these protocols with E gene amplification for detection and RdRp gene amplification for confirmation^7^. The RdRp gene primers and probe were 100% identical to the biding sites of 100% (388/388) available RdRp gene sequences of SERS-CoV-2 deposited in GenBank. The E gene qPCR primers and probe were identical to the biding site of 99.7% (375/376). The E gene probe had 1 bp mismatch with only 1 E gene sequence MT039890 (SERS-CoV-2/SNU01 from Korea).

Hong Kong’s qPCR protocol consisted of Assay 1 targeting orf1b-nsp14 and Assay 2 targeting the N gene. The orf1b primers and probe had perfect match with 99.5% (373/375) of the SARS-CoV-2 sequences. The reverse primer had 1 base different from sequences MT163712 (SARS-CoV-2/human/IRN/mehr1/2020 from Iran) and MT276327 (SARS-CoV-2 isolate SARS-CoV-2/h uman/USA/GA_2742/2020). The N gene primers and probe had perfect match with the biding sites of 99.5% (383/385) of the SARS-CoV-2 sequences with completed target sequences. The probe had 1 base different from sequences MT263458 (SARS-CoV-2/human/USA/WA-UW379/2020) and MT263435 (SARS-CoV-2/human/USA/WA-UW355/2020).

The Department of Medical Sciences, Ministry of Public Health, Thailand used an assay targeting the N gene. The primer and probe sequences are identical to all but one of 386 SARS-CoV-2 sequences with available in GenBank. Sequence MT184913 (2019-nCoV/USA-CruiseA-26/2020) had an ambiguous code “Y” (for C/T) which has 50% possibility aligning with “C” of the reverse primer.

## Discussion

After looking at available primer and probe sequences, it appears as though at present (April 2020) most sequences for the qRT-PCR detection of SARS-CoV-2 largely match the current reported genomic sequences. The three bp disparity between the Chinese CDC N gene primers and strains isolated from five different countries including Spain, Israel, India, Peru and America highlights the importance to continue monitoring the performance of the PCR protocol and re-evaluate them at the least in silico when more sequences become available.

Given the challenges in obtaining clinical samples from which virus may be detected and variations between laboratories, even single bp mismatches represent errors of 5% in the primer sequence and depending on their location and the nature of the substitution, these could affect performance of the reaction and yield false negative results^8^. Indeed, a single base pair mismatch was shown to reduce sensitivity in a qRT-PCR test kit distributed during the 2009 H1N1 pandemic^9^. This evaluation was on primer and probe sequences only. The other characteristics, e.g. annealing temperature, primer dimer and hair pin forming possibility of the primers and probes were not evaluated. Given substantial contributions to qRT-PCR efficiency from these factors, sequence changes represent a point of importance when considering diagnostic tests.

SARS-CoV-2 likely mutates at a rate similar to the first SARS coronavirus, over 60 mutations per genome per year^10^. It is important then, that continued sharing of sequencing data becomes increasingly important as the pandemic currently shows few signs of slowing down and could remain in circulation for a number of years. Already there are over 8,000 sequences in the GISAID hCoV-19 database (as of mid-April 2020) that demonstrate considerable phylogenetic diversity^11–13^. It is important to note as well, that sequencing errors could also have played a role in the incorrect sequences, but this is unlikely given a conserved mutation in the aforementioned 13 isolates.

In the current emergency, the low sensitivity of qRT-PCR implies that many COVID-19 patients may not be identified and may not receive appropriate treatment in time; such patients constitute a risk for infecting a larger population given the highly contagious nature of the virus. Particularly as countries around the world are rushing to acquire sufficient testing capacity and develop containment strategies^14^, it will be particularly important to ensure that design of primers and probes stays current with the evolving SARS-CoV-2 genetic sequence.

## Materials and Methods

### Study Identification and Selection

A systematic search was carried out in three major electronic databases (PubMed, Embase and Cochrane Library) to identify published studies examining the diagnosis, therapeutic drugs and vaccines for Severe Acute Respiratory Syndrome (SARS), Middle East Respiratory Syndrome (MERS) and the 2019 novel coronavirus (2019-nCoV), in accordance with the Preferred Reporting Items for Systematic Reviews and Meta-Analyses (PRISMA) guidelines.

As of April 6, 2020, a total of 447 SARS-CoV-2 (COVID-19 pathogen) sequences were available from the GenBank, including 375 complete or close to complete genome with greater than 29,161 bp (hereafter name as full genome), 72 partial genome sequences with 87bp to 1,411 bp.

All 375 full-length sequences were downloaded and used to create a custom SARS-CoV-2 genome database using a bioinformatic software Geneious version 11.1 (Biomatters, Auckland, New Zealand). PCR protocols were collected from publication of WHO, US CDC and other literatures. Primers and probes of each protocols were blast searched against the custom SARS-CoV-2 genome database, and or analyzed with multiple sequence alignment using Geneious version 11.1.

## Data Availability

All data has been acquired in accordance with the Preferred Reporting Items for Systematic Reviews and Meta-Analyses (PRISMA) guidelines and is freely available on GenBank and from the World Health Organization website.

https://www.who.int/docs/default-source/coronaviruse/whoinhouseassays.pdf?sfvrsn=de3a76aa_2

https://www.ncbi.nlm.nih.gov/genbank/

## References

1. Pang, J., Wang, M. X., Ang, I. Y. H., Tan, S. H. X., Lewis, R. F., Chen, J. I.-P., Gutierrez, R. A., Gwee, S. X. W., Chua, P. E. Y., Yang, Q., Ng, X. Y., Yap, R. K. S., Tan, H. Y., Teo, Y. Y., Tan, C. C., Cook, A. R., Yap, J. C.-H. & Hsu, L. Y. Potential Rapid Diagnostics, Vaccine and Therapeutics for 2019 Novel Coronavirus (2019-nCoV): A Systematic Review. J. Clin. Med. 9, 623 (2020).

2. Sanders, R., Mason, D. J., Foy, C. A. & Huggett, J. F. Considerations for accurate gene expression measurement by reverse transcription quantitative PCR when analysing clinical samples. Anal. Bioanal. Chem. 406, 6471–6483 (2014).

3. Murphy, J. & Bustin, S. A. Reliability of real-time reverse-transcription PCR in clinical diagnostics: gold standard or substandard? Expert Rev. Mol. Diagn. 9, 187–197 (2009).

4. Lu, R., Zhao, X., Li, J., Niu, P., Yang, B., Wu, H., Wang, W., Song, H., Huang, B., Zhu, N., Bi, Y., Ma, X., Zhan, F., Wang, L., Hu, T., Zhou, H., Hu, Z., Zhou, W., Zhao, L., Chen, J., Meng, Y., Wang, J., Lin, Y., Yuan, J., Xie, Z., Ma, J., Liu, W. J., Wang, D., Xu, W., Holmes, E. C., Gao, G. F., Wu, G., Chen, W., Shi, W. & Tan, W. Genomic characterisation and epidemiology of 2019 novel coronavirus: implications for virus origins and receptor binding. The Lancet 395, 565–574 (2020).

5. Wang, W., Xu, Y., Gao, R., Lu, R., Han, K., Wu, G. & Tan, W. Detection of SARS-CoV-2 in Different Types of Clinical Specimens. JAMA (2020). doi:10.1001/jama.2020.3786

6. SARS nCoV-2 PCR Protocols. at <https://www.who.int/docs/default-source/coronaviruse/whoinhouseassays.pdf?sfvrsn=de3a76aa_2>

7. Coronavirus Disease 2019 (COVID-19) Testing. At <https://www.publichealthontario.ca/en/laboratory-services/test-information-index/wuhan-novel-coronavirus>

8. Stadhouders, R., Pas, S. D., Anber, J., Voermans, J., Mes, T. H. M. & Schutten, M. The Effect of Primer-Template Mismatches on the Detection and Quantification of Nucleic Acids Using the 5’ Nuclease Assay. J. Mol. Diagn. 12, 109–117 (2010).

9. Klungthong, C., Chinnawirotpisan, P., Hussem, K., Phonpakobsin, T., Manasatienkij, W., Ajariyakhajorn, C., Rungrojcharoenkit, K., Gibbons, R. V. & Jarman, R. G. The impact of primer and probe-template mismatches on the sensitivity of pandemic influenza A/H1N1/2009 virus detection by real-time RT-PCR. J. Clin. Virol. 48, 91–95 (2010).

10. Vega, V. B., Ruan, Y., Liu, J., Lee, W. H., Wei, C. L., Se-Thoe, S. Y., Tang, K. F., Zhang, T., Kolatkar, P. R., Ooi, E. E., Ling, A. E., Stanton, L. W., Long, P. M. & Liu, E. T. Mutational dynamics of the SARS coronavirus in cell culture and human populations isolated in 2003. BMC Infect. Dis. 4, 32 (2004).

11. Hadfield, J., Megill, C., Bell, S. M., Huddleston, J., Potter, B., Callender, C., Sagulenko, P., Bedford, T. & Neher, R. A. Nextstrain: real-time tracking of pathogen evolution. Bioinformatics 34, 4121–4123 (2018).

12. Shu, Y. & McCauley, J. GISAID: Global initiative on sharing all influenza data – from vision to reality. Eurosurveillance 22, 30494 (2017).

13. Elbe, S. & Buckland-Merrett, G. Data, disease and diplomacy: GISAID’s innovative contribution to global health: Data, Disease and Diplomacy. Glob. Chall. 1, 33–46 (2017).

14. Fisher, D. & Wilder-Smith, A. The global community needs to swiftly ramp up the response to contain COVID-19. The Lancet 395, 1109–1110 (2020).

